# The relationships between MASLD, extrahepatic multimorbidity and all-cause mortality in UK Biobank cohort

**DOI:** 10.1101/2025.03.09.25323328

**Authors:** Qi Feng, Chioma N Izzi-Engbeaya, Andrea D. Branch, Benjamin H Mullish, Pinelopi Manousou, Mark Woodward

**Affiliations:** The George Institute for Global Health (UK), School of Public Health, Faculty of Medicine, Imperial College London, London, UK; Section of Investigative Medicine and Endocrinology, Department of Metabolism, Digestion and Reproduction, Faculty of Medicine, Imperial College London, UK; Department of Medicine, Division of Liver Diseases, the Icahn School of Medicine at Mount Sinai, New York, NY, USA; Division of Digestive Diseases, Department of Metabolism, Digestion and Reproduction, Faculty of Medicine, Imperial College London, London, UK; Department of Hepatology, St Mary’s Hospital, Imperial College Healthcare NHS Trust, London, UK; The George Institute for Global Health (Australia), University of South New Wales, Sydney, Australia

**Keywords:** MASLD, NAFLD, multimorbidity, all-cause mortality, UK Biobank, long-term conditions, sexual dimorphism

## Abstract

**Background & Aims:** This study aimed to estimate the impact of metabolic dysfunction-associated steatotic liver disease (MASLD), with and without multimorbidity, on all-cause mortality.

**Methods:** We analysed data from the UK Biobank. MASLD was identified as a fatty liver index (FLI) ≥ 60 and presence of cardiometabolic risk factors. Multimorbidity was defined as ≥2 of the long-term conditions (LTCs) in a pre-specified list of 47 extrahepatic conditions. Hazard ratios (HRs) from adjusted Cox models quantified the association between MASLD, multimorbidity and all-cause mortality.

**Results:** Of the 438,840 participants, 131,020 (29.9%) had MASLD at baseline. The participants with MASLD at baseline had a higher prevalence of multimorbidity than those without (21.3% vs. 14.4%). In addition to cardiometabolic risk factors, MASLD was strongly associated with several LTCs, particularly metabolic, cardiovascular, cancers, kidney, mental/behavioural, and respiratory diseases. During a median follow-up of 13 years, MASLD was associated with higher mortality (HR 1.16 (95%CI: 1.13, 1.19)), with stronger associations in females and in those with low LTC counts (≤3 LTCs). Each additional LTC at baseline was associated with 30% and 38% higher mortality in MASLD (HR 1.30 (1.29, 1.32)) and non-MASLD (HR 1.38 (1.37, 1.40)) populations, respectively. Among the 47 LTCs, 16 were associated with increased mortality in people with MASLD.

**Conclusion:** Those with MASLD exhibited a higher prevalence of extrahepatic multimorbidity and a 16% higher rate of mortality than those without, underscoring the impact of liver steatosis on mortality and highlighting the need to target LTCs to improve outcomes and reduce healthcare burdens.

## Introduction

Metabolic dysfunction-associated steatotic liver disease (MASLD), the replacement term for non-alcoholic fatty liver disease (NAFLD), is defined as the presence of liver steatosis along with one or more case-defining cardiometabolic risk factors (obesity, diabetes, hypertension and dyslipidaemia), in the absence of excessive alcohol consumption or other established chronic liver diseases [1]. MASLD consists of a spectrum of hepatic conditions, ranging from simple steatosis to steatohepatitis, and can progress to fibrosis, cirrhosis, and hepatocellular carcinoma [1,2]. MASLD, the most common liver condition, has emerged as a significant public health concern globally, with a prevalence of 32.4% [2,3]. The high prevalence of MASLD is associated with an increased risk of hepatic morbidity and mortality, as well as a substantial economic burden [4,5].

Multimorbidity is defined here as the coexistence of two or more long term conditions (LTCs) that are on a specific pre-defined list [6]. Multimorbidity increases the severity and progression rate of many adverse health outcomes, such as cognitive decline [7], cancers [8], cardiovascular disease [9], reduced health-related quality of life [10], and mortality [11]. While the burden of multimorbidity has been studied in the general population [8,12], and in specific groups, such as people with diabetes [13] and with stroke [14], its implications in individuals with MASLD have not been comprehensively studied.

MASLD is considered to be the hepatic manifestation of metabolic syndrome [15] and is associated with diabetes [16], cardiovascular disease [17], extrahepatic cancers [18] and chronic kidney disease [19]. Consequently, individuals with MASLD are likely to be at increased risk for extrahepatic multimorbidity. However, previous research investigating multimorbidity in MASLD patients often focused on a narrow range of conditions [20,21]. For example, an Italian study described multimorbidity in 95 patients with diabetes and NAFLD but only considered 16 LTCs [22]. A study in Israel examined 200,000 patients with NAFLD but included only 14 LTCs, half of which were metabolic disorders [23]. Therefore, a formal investigation of the relationship between MASLD and multimorbidity defined using a more extensive list of LTCs is warranted.

In the UK, liver-related mortality has increased four-fold since the 1970s, positioning liver disease as the third most common cause of premature death [24]. MASLD has been an increasingly important contributor to liver-related mortality. Although it is a distant second to alcohol-related liver disease, MASLD surpasses viral hepatitis, autoimmune or metabolic liver diseases and other causes in the UK [25]. MASLD has become one of the most common indications for liver transplantation [26]. To understand the clinical consequences of metabolic dysfunction, it is crucial to understand MASLD, extrahepatic multimorbidity and their interactions with health outcomes.

Accordingly, we aimed to describe the prevalence of multimorbidity, in individuals with MASLD, and to quantify the impact of MASLD, in conjunction with the degree of multimorbidity, on all-cause mortality in a large UK-based study.

## Methods

We used data from the UK Biobank, a population-based cohort of half million participants aged between 40 and 70 years old recruited between 2006 and 2010 [27]. At baseline assessment, data were collected on sociodemographic characteristics, lifestyle, health status, medication use, environmental factors, anthropometry and physical measures. Biological samples including blood, saliva and urine were collected. Participants were prospectively followed up via linkage to national death registries, cancer registries, and hospitalisation data [27]. UK Biobank was approved by the North-West Multi-centre Research Ethics Committee, the National Information Governance Board for Health and Social Care in England and Wales, and the Community Health Index Advisory Group in Scotland. All participants provided informed consent.

We used the definition proposed by Rinella M. et al. to identify individuals with MASLD, which consists of liver steatosis, at least one cardiometabolic risk factor and low alcohol consumption (< 20 (female) /30 (male) g/day) [28]. We used fatty liver index (FLI) ≥ 60% as an indicator of liver steatosis. FLI is a score representing hepatic steatosis levels, calculated from the body mass index (BMI), waist circumference, triglycerides, and gamma-glutamyl transferase (GGT) [29]. The cutoff value of 60% has been used previously to define steatosis [4,5]. The accuracy of FLI in identifying individuals with steatosis has been validated [5]. We further identified individuals with any of the following cardiometabolic risk factors: obesity (body mass index (BMI) ≥ 25 kg/m ² AND/OR waist circumference >94 cm (male) (>90 cm (female))), diabetes (glycated haemoglobin (HbA1c)≥48mmol/mol AND/OR diagnosis of type 2 diabetes AND/OR on treatment for type 2 diabetes), hypertension (systolic blood pressure (BP) ≥ 130 AND/OR diastolic BP ≥ 85 mmHg AND/OR on antihypertensive drug treatment or diagnosis of hypertension), high triglycerides (TG) (plasma TG ≥1.70 mmol/L AND/OR on lipid lowering treatment), and low high-density lipoprotein (HDL) cholesterol (HDL-cholesterol ≤ 1.0 mmol/L (male) (≤ 1.3 mmol/L (female)) AND/OR on lipid lowering treatment) [28].

We excluded women who were pregnant at baseline, and people who had missing data for calculating FLI or defining MASLD. We also excluded people with alcohol related liver disease (ALD), metabolic dysfunction and alcohol related liver disease (MetALD), or other chronic liver conditions (including viral hepatitis, liver fibrosis, liver cirrhosis, hepatocellular carcinoma, hemochromatosis, Wilson’s disease, biliary cirrhosis, autoimmune hepatitis, primary sclerosing cholangitis, toxic liver disease and Budd-Chiari syndrome). For ALD and MetALD, we used the definition proposed by Rinella M. et al. [28]., whereas other chronic liver disease was ascertained via self-report and hospitalisation data; the code lists for these conditions are presented in the supplementary methods.

### Exposure

We used the self-report data and hospitalisation records on and before the baseline visit for phenotyping baseline multimorbidity. We defined multimorbidity as having two or more LTCs from a pre-specified list. We based our list of LTCs from those identified in a three-round Delphi study of 25 public participants and 150 healthcare professionals (including clinicians, researchers and policy makers) in 2021 [30]. The criteria for including these LTCs included their impacts on risk of death, quality of life, frailty, physical disability, mental health and treatment burden. Among the conditions identified in this Delphi study, we combined solid organ cancers, metastatic cancers, melanoma, and treated cancer requiring surveillance, into one broad solid organ cancer category, and removed post-acute Covid-19, chronic Lyme disease, and two liver conditions (hepatocellular carcinoma and chronic liver disease). Since MASLD is our index disease, we excluded MASLD and its five associated cardiometabolic risk factors (obesity, hypertension, diabetes, high TG and low HDL-cholesterol) from the list. After these modifications, this study considered 47 LTCs for multimorbidity (supplementary methods). These LTCs covered extrahepatic cancers, cardiovascular, metabolic, endocrinological, respiratory, digestive, renal, mental/behavioural and congenital conditions.

### Outcome

Our outcome was all-cause mortality within the follow-up period of the study, confirmed via death registry records. Participants were censored at the date of death or the last date of follow-up (30 November 2022), whichever occurred first.

#### Covariates

A participant’s region was determined by the location of the assessment centre they attended at baseline. Ethnicity was classified into White, Asian, Black, and mixed/others. The Townsend Deprivation Index is a postcode-derived measure used to designate socioeconomic status. Educational attainment was categorised as below secondary, lower secondary, upper secondary, vocational training, and higher education. Lifestyle factors considered were self-reported current smoking (current, previous and never smoker), alcohol consumption, and physical activity level. Alcohol consumption was assessed via self-reported weekly or monthly intake of red wine, white wine, champagne, beer, spirits, fortified wine and other alcoholic drinks; the consumption was converted to standard UK alcohol units and further to alcohol grams and summed up to derive the average daily alcohol consumption (g/d) [31]. Physical activity level was measured with the International Physical Activity Questionnaire and centrally processed by the UK Biobank; individuals were categorized into low, moderate and high levels, based on the frequency, duration and intensity of their physical activities. Systolic and diastolic BP were measured by trained staff twice within a few minutes apart and the averages of the two readings were used in analyses. Blood biochemistry markers were measured at a central laboratory by the UK Biobank using the blood sample collected at baseline, including TG, HDL-cholesterol, low density lipoprotein cholesterol, glucose, HbA1c, and haemoglobin levels. For all the categorical covariates, answers of “unknown”, “do not know”, “prefer not to say” were combined into one “unknown” category.

### Statistical analysis

The baseline characteristics were summarised using mean with standard deviation or median with interquartile interval, as appropriate, and frequency with percentage, stratified by MASLD status and the number of LTCs (0, 1 and ≥ 2 LTCs).

We used bar charts to demonstrate the distribution of the number of LTCs stratified by MASLD status, sex and age groups (< 50, 50-59, ≥ 60 years). We calculated the prevalence (per 1000) for each LTC and ranked these factors in people with and without MASLD separately. We fitted logistic regression models, adjusted for sex, age, education and Townsend Deprivation Index (in fifths), to estimate prevalence odds ratios (ORs) and their 95% confidence intervals (CIs) relating MASLD status to each LTC cross-sectionally; p values were adjusted for multiple testing using the false discovery rate (FDR) method [32].

Cox proportional hazard regression models were used to assess the association between having MASLD and all-cause mortality, expressed as hazard ratio (HR) with 95% confidence interval (CI). Models were stratified by region and age group, and adjusted for sex, ethnicity, education, Townsend Deprivation Index (in fifths), physical activity level, smoking status, alcohol consumption and LTC counts. The proportional hazard assumption was examined by scaled Schoenfeld residuals, and no evidence was observed for its violation. This analysis was repeated in people with different LTC counts (0, 1, 2, 3, 4, ≥5), thus enabling us to assess the effect of additionally having MASLD on all-cause mortality, across a range of LTC counts. We also evaluated the association between the number of LTCs and all-cause mortality with similarly adjusted Cox models. The number of LTCs was fitted as a categorical (0, 1, 2, 3, 4, and ≥ 5 LTCs) and as a continuous (per each additional LTC) variable in separate models. We assessed the association of multimorbidity separately in MASLD and non-MASLD cohorts, and compared the two HRs using the ratio of hazard ratios (RHR), with its 95% CI [33]. In sensitivity analysis, we (1) adjusted the Cox models additionally for the five cardiometabolic risk factors, and (2) excluded the first two years of follow-up to correct for reverse causation. The association between LTC numbers and all-cause mortality was also assessed in subgroups stratified by age group, Townsend Deprivation Index (in fifths), education, smoking status, physical activity and BMI levels.

We assessed the association between cardiometabolic risk factors and mortality, using people without the risk factor under examination as the reference group, in people with and with MASLD separately. We assessed the associations between specifically having any of the 47 LTCs and all-cause mortality, using people without any LTC as the reference group, in people with MASLD.

All analyses were conducted in R software and we applied a correction for multiple comparisons [32]. This study is compliant with the Strobe checklist for cohort study.

## Results

### Participants and baseline characteristics

Among 438,840 eligible individuals (mean age 56.5 (SD 8.1) years, 42.2% males), we identified 131,020 (29.9%) individuals with MASLD (mean age 57.4 (SD 7.9), 58.2% males)) (figure 1). MASLD prevalence was higher in males (41.2%) than females (21.6%) and in older people (24.5%, 29.7% and 32.0% for people < 50, 50-59, ≥ 60 years, respectively).

**Figure 1:**
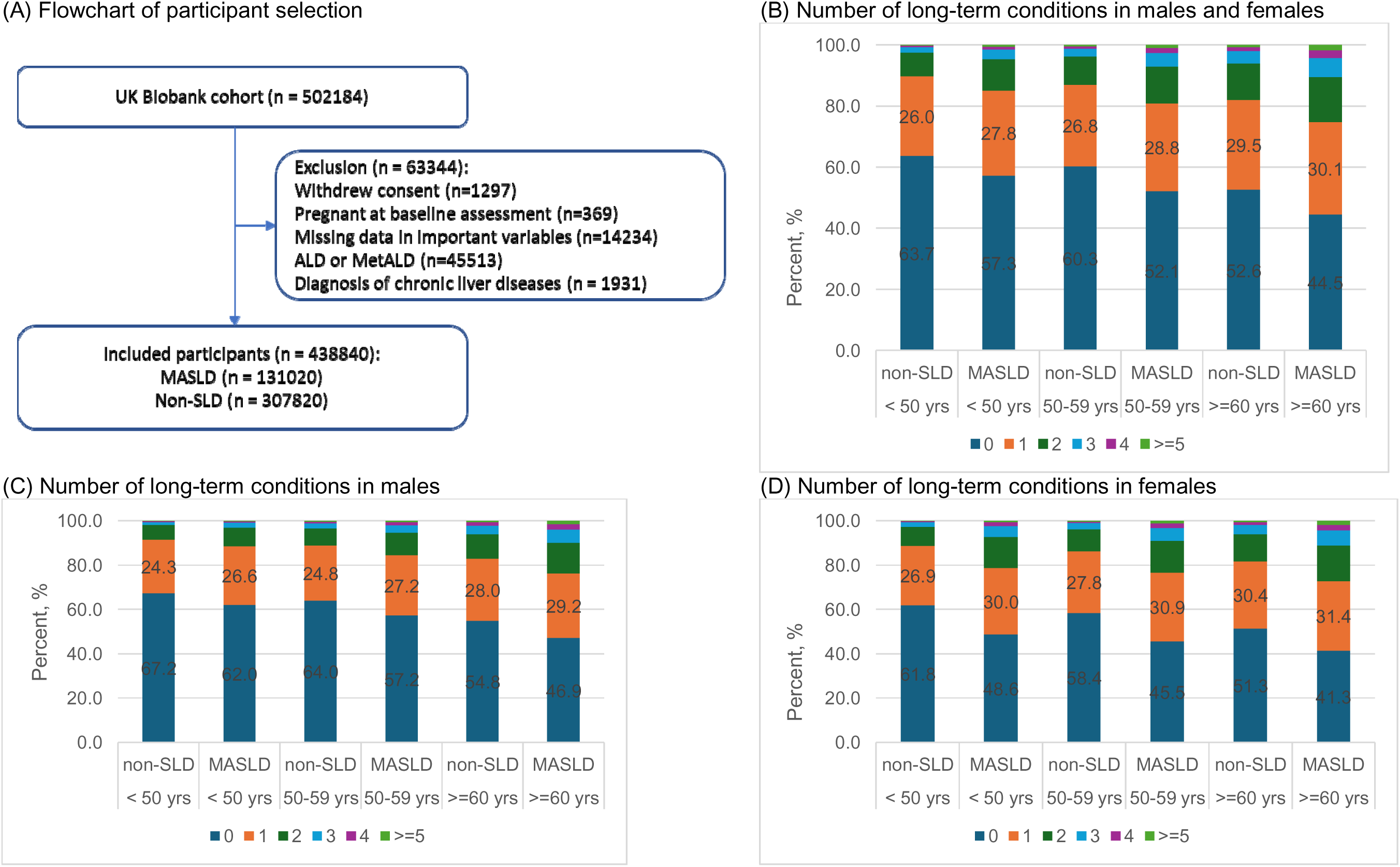
The flowchart of participants selection and the number of long-term conditions in people with and without MASLD, stratified by sex and age group. (A) Flowchart of participant selection. (B) Number of long-term conditions in males and females. (C) Number of long-term conditions in males. (D) Number of long-term conditions in females. SLD: steatotic liver disease. MASLD: metabolic dysfunction-associated steatotic liver disease. ALD: alcohol related liver disease. MetALD: metabolic dysfunction and alcohol related liver disease.

Overall, 28.2% of participants had one of the designated LTC, and 16.4% had ≥ 2 LTCs. The prevalence of having ≥2 LTCs was slightly higher in females (17.2%) than males (15.4%), in those with MASLD (21.3%) than in people without (14.4%), and increased with age (11.5%, 14.8% and 20.4% for those <50, 50-59, and ≥ 60 years, respectively) (figure 1).

Individuals with MASLD were more likely, than those without, to be of older age, male, more deprived, less educated, less physically active, and to have higher BMI, waist circumference, blood pressures, TG, glucose, HbA1c, and to have lower HDL (Table 1). The three most prevalent cardiometabolic risk factors were obesity (98.9%), hypertension (80.3%) and high TG (72.6%) for people with MASLD, and hypertension (61.2%), obesity (55.9%) and low HDL-cholesterol (30.1%) for people without MASLD. Among people with MASLD, 3.2%, 19.2%, 35.2%, 29.5% and 12.9% had one, two, three, four and five cardiometabolic risk factors, respectively. Among people without MASLD, 12.7%, 28.2%, 31.6%, 18.2%, 7.5% and 1.8% had zero, one, two, three, four and five cardiometabolic risk factors, respectively. People with more LTCs were more likely to be older, more socioeconomically deprived, less educated, less physically active, and exhibited higher BMI, waist circumference, and HbA1c. As expected, each of the five MASLD-defining cardiometabolic risk factors was more prevalent in the MASLD cohort than the non-MASLD cohort. Supplementary table 1 provides more details about baseline characteristics.

**Table 1:** Baseline characteristics of participants with and without MASLD, stratified by number of long-term conditions.

|  | No long-term condition |  | 1 long-term condition |  | 2 or more long-term conditions |  | Total<br>n = 438840 |
| --- | --- | --- | --- | --- | --- | --- | --- |
|  | Non-SLD | MASLD | Non-SLD | MASLD | Non-SLD | MASLD |  |
|  | n = 178357 | n = 64823 | n = 85262 | n = 38312 | n = 44201 | n = 27885 |  |
| Sex, male | 66167 (37.1%) | 40767 (62.9%) | 28414 (33.3%) | 21333 (55.7%) | 14374 (32.5%) | 14175 (50.8%) | 185230 (42.2%) |
| Age, years | 55.4 (8.2) | 56.6 (7.9) | 56.6 (8.2) | 57.7 (7.8) | 58.1 (8.0) | 59.1 (7.5) | 56.5 (8.1) |
| Townsend Deprivation Index |  |  |  |  |  |  |  |
| 1st fifth (least deprived) | 38336 (21.5%) | 12558 (19.4%) | 18009 (21.1%) | 6982 (18.2%) | 8355 (18.9%) | 4086 (14.7%) | 88326 (20.1%) |
| 5th fifth (most deprived) | 31181 (17.5%) | 13685 (21.1%) | 15488 (18.2%) | 8918 (23.3%) | 9996 (22.6%) | 8320 (29.8%) | 87588 (20.0%) |
| Education, higher education | 66587 (37.3%) | 18240 (28.1%) | 29978 (35.2%) | 10026 (26.2%) | 13307 (30.1%) | 5942 (21.3%) | 144080 (32.8%) |
| Ethnicity, White | 167842 (94.1%) | 59845 (92.3%) | 80866 (94.8%) | 35899 (93.7%) | 41951 (94.9%) | 26175 (93.9%) | 412578 (94.0%) |
| Smoking, never | 107463 (60.3%) | 36154 (55.8%) | 49133 (57.6%) | 20163 (52.6%) | 22892 (51.8%) | 12829 (46.0%) | 248634 (56.7%) |
| Alcohol drinking, g/d | 14.4 (16.7) | 9.2 (9.1) | 13.4 (16.5) | 8.1 (8.8) | 12.4 (18.8) | 6.6 (8.4) | 12.2 (15.2) |
| Physical activity, high | 63798 (35.8%) | 17614 (27.2%) | 28765 (33.7%) | 9690 (25.3%) | 13640 (30.9%) | 6102 (21.9%) | 139609 (31.8%) |
| ALT, log<sub>10</sub> U/L | 1.2 (0.3) | 1.4 (0.2) | 1.2 (0.3) | 1.4 (0.2) | 1.2 (0.3) | 1.4 (0.2) | 1.3 (0.3) |
| AST, log<sub>10</sub> U/L | 1.3 (0.2) | 1.4 (0.1) | 1.3 (0.2) | 1.4 (0.1) | 1.3 (0.3) | 1.4 (0.2) | 1.3 (0.2) |
| FIB4 score | -- | 1.3 (1.3) | -- | 1.3 (0.7) | -- | 1.4 (6.3) | 1.3 (2.0) |
| BMI, kg/m2 | 25.0 (3.0) | 31.4 (4.4) | 25.1 (3.1) | 31.9 (4.6) | 25.3 (3.3) | 32.7 (5.1) | 27.1 (4.7) |
| Waist circumference, cm | 82.9 (9.4) | 101.9 (9.7) | 83.3 (9.4) | 102.6 (10.2) | 84.0 (9.7) | 104.4 (11.1) | 89.0 (13.1) |
| Systolic blood pressure, mmHg | 135.2 (18.6) | 142.5 (17.4) | 135.1 (18.6) | 141.5 (17.4) | 135.3 (18.9) | 139.8 (17.7) | 137.1 (18.6) |
| Diastolic blood pressure, mmHg | 80.5 (9.8) | 86.0 (9.5) | 80.2 (9.8) | 84.9 (9.6) | 79.7 (10.0) | 83.3 (10.1) | 81.7 (10.0) |
| Triglyceride, mmol/L | 1.1 [0.7] | 2.2 [1.3] | 1.2 [0.7] | 2.1 [1.3] | 1.2 [0.8] | 2.1 [1.3] | 1.4 [1.0] |
| HDL-cholesterol, mmol/L | 1.4 (0.6) | 1.1 (0.4) | 1.4 (0.6) | 1.1 (0.4) | 1.3 (0.6) | 1.1 (0.4) | 1.3 (0.6) |
| HbA1c, mmol/mol | 33.8 (6.5) | 36.5 (9.5) | 34.1 (6.7) | 37.2 (10.0) | 34.8 (7.8) | 38.7 (11.5) | 35.0 (8.1) |
| Hypertension | 108041 (60.6%) | 52652 (81.2%) | 52413 (61.5%) | 30724 (80.2%) | 27943 (63.2%) | 21801 (78.2%) | 293574 (66.9%) |
| Obesity | 96588 (54.2%) | 63985 (98.7%) | 48852 (57.3%) | 37914 (99.0%) | 26674 (60.3%) | 27654 (99.2%) | 301667 (68.7%) |
| Diabetes | 19668 (11.0%) | 17394 (26.8%) | 10935 (12.8%) | 12112 (31.6%) | 7830 (17.7%) | 11403 (40.9%) | 79342 (18.1%) |
| High triglyceride | 41605 (23.3%) | 46871 (72.3%) | 22447 (26.3%) | 27820 (72.6%) | 13868 (31.4%) | 20425 (73.2%) | 173036 (39.4%) |
| Low HDL-cholesterol | 49354 (27.7%) | 27731 (42.8%) | 26658 (31.3%) | 18137 (47.3%) | 16544 (37.4%) | 15289 (54.8%) | 153713 (35.0%) |

HbA1c: glycated hemoglobin. HDL: high density lipoprotein. LDL: low density lipoprotein. Number(number): mean(SD).
Number(percent%): frequency(percent). Number[number]: median[interquartile range].

### Prevalence of cardiometabolic risk factors and LTCs

The crude prevalence of each condition, stratified by MASLD and sex, is shown in supplementary table 2.

Overall, the top six most prevalent LTCs were similar in people with and without MASLD (supplementary figure 1), although rankings varied. The top six conditions were asthma, chronic respiratory disease (excluding asthma and COPD), thyroid disorder, depression, extrahepatic solid organ cancers and osteoporosis in females without MASLD; asthma, thyroid disorder, chronic respiratory disease, depression, extrahepatic solid organ cancer and ischemic heart disease (IHD) in females with MASLD; chronic respiratory disease, asthma, IHD, extrahepatic solid organ cancer, depression and arrythmia in males without MASLD; and chronic respiratory disease, asthma, IHD, depression, extrahepatic solid organ cancer and arrythmia in males with MASLD.

After adjusting for sex, age, education and Townsend Deprivation Index, individuals with MASLD were more likely to have 32 of the 47 LTCs, with the highest ORs for gout (2.83 (2.42, 3.32)), heart failure (2.24 (2.03, 2.46)), post traumatic stress disorder (PTSD) (2.13 (1.68, 2.68)), osteoarthritis (1.98 (1.91, 2.06)), and bipolar disorder (1.90 (1.71, 2.12)). Overall, metabolic, mental/behavioural, cardiovascular, and renal conditions were more prevalent in MASLD patients, while substance use disorder (0.83 (0.77, 0.89)), osteoporosis (0.63 (0.60, 0.67)) and eating disorder (0.42 (0.30, 0.59)) were less common in people with MASLD than in people without. Most ORs were higher for females than for males; for example, the ORs for gout were 4.76 (3.12, 7.25) and 2.57 (2.16, 3.04), for stroke 1.66 (1.54, 1.80) and 1.41 (1.32, 1.51), for IHD 2.13 (2.03, 2.24) and 1.71 (1.65, 1.77), for COPD 1.34 (1.25, 1.43) and 1.16 (1.08, 1.24), for transient ischemic attack (TIA) 1.52 (1.24, 1.85) and 1.26 (1.06, 1.49), for females and males, respectively. Some LTCs were significantly associated with MASLD in females, but not in males, such as multiple sclerosis (1.18 (1.03, 1.36) vs. 1.01 (0.82, 1.23)) and extrahepatic solid organ cancers (1.09 (1.05, 1.14) vs. 1.05 (0.99, 1.10)) (table 2).

**Table 2:**
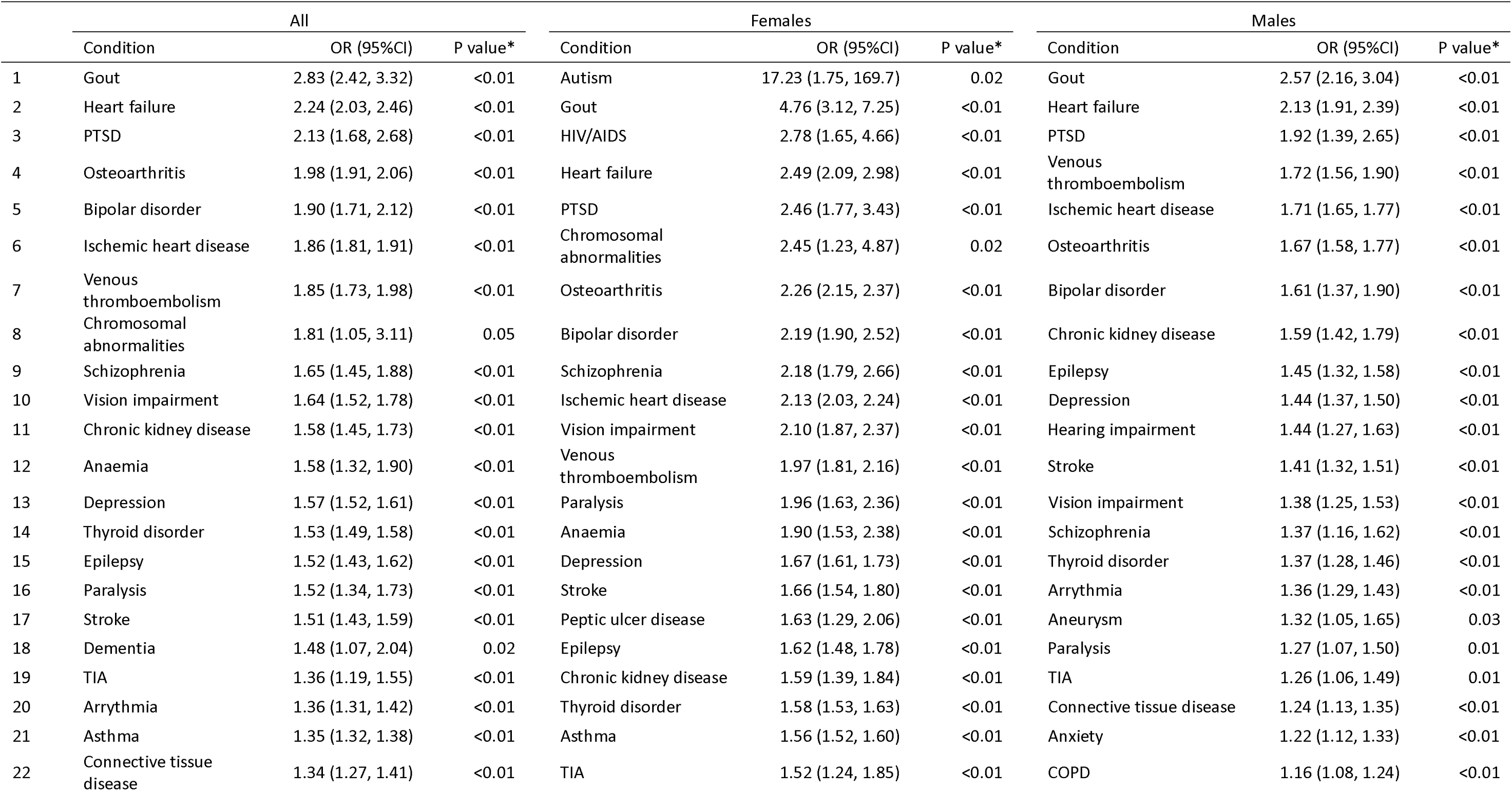
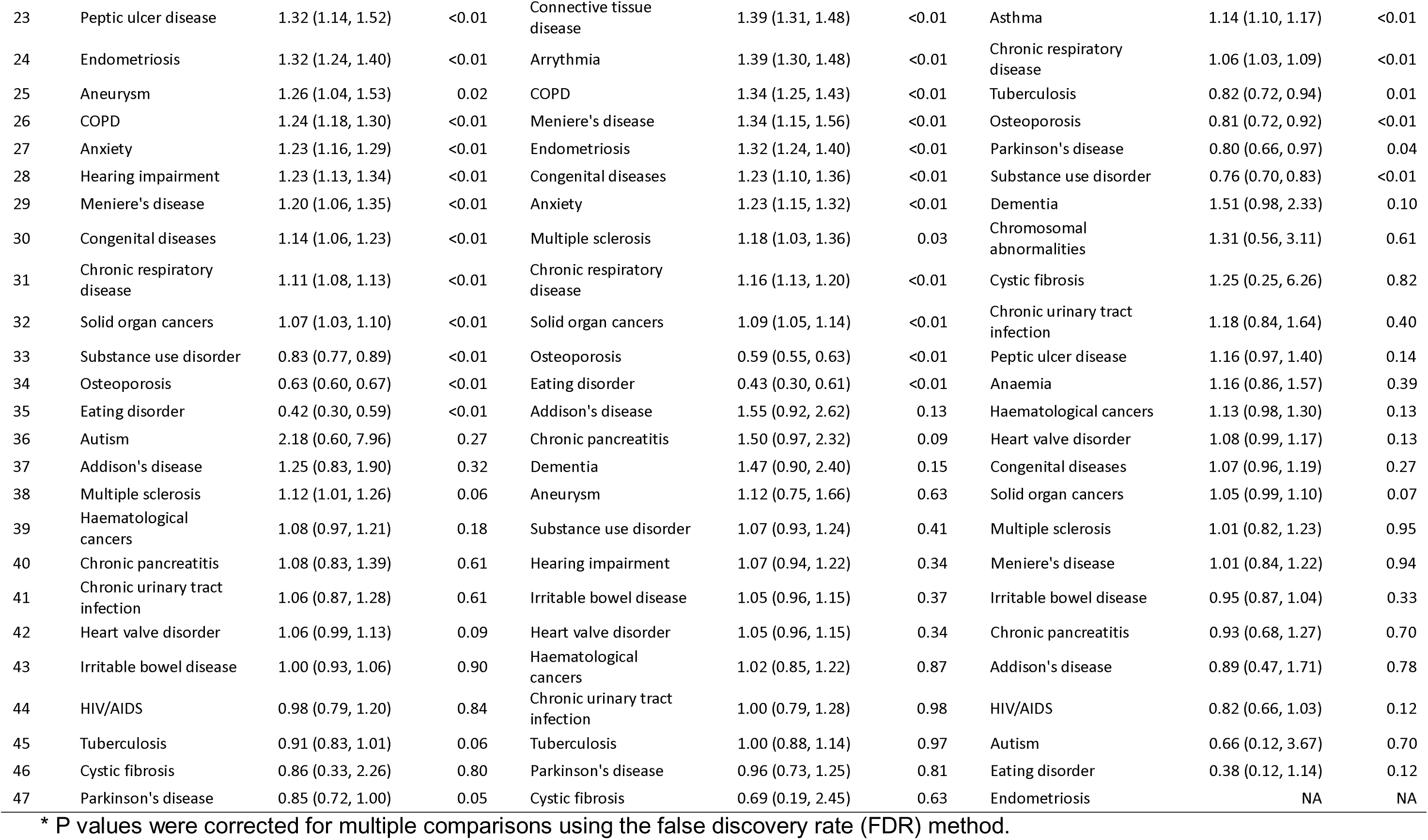
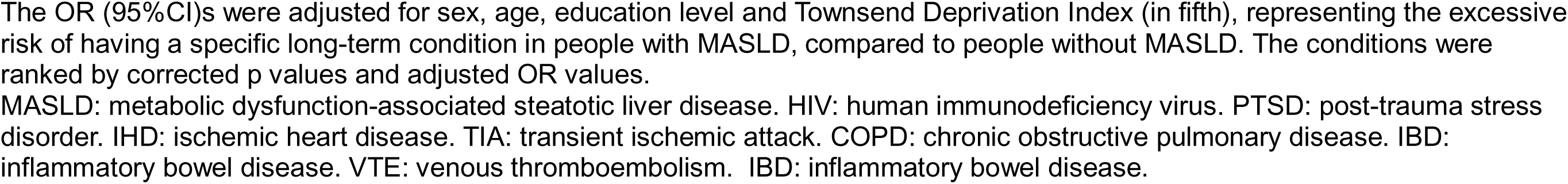
Adjusted prevalence odds ratios (95% confidence intervals) comparing MASLD to non-MASLD on long-term conditions at baseline, by sex.

### MASLD, Multimorbidity and all-cause mortality

During a median follow up of 13 years, 36,664 deaths occurred, with 14,910 among people with MASLD. The major causes of death were cancers, circulatory, respiratory, nervous system, and digestive diseases, in both MASLD and non-MASLD cohorts, with cancers, circulatory, respiratory and nervous diseases responsible for 80% of deaths.

MASLD was positively, but modestly, associated with increased all-cause mortality after adjusting for the number of LTCs (HR 1.16 (1.13, 1.19)), and had a greater impact on females (HR 1.25 (1.20, 1.29)) than on males (1.10 (1.07, 1.13)). When looking at subgroups stratified by the number of LTCs, having MASLD was associated with increased mortality only when the LTC counts were low (< 3 in females and <2 in males) (table 3).

**Table 3:** Hazard ratios (95% confidence interval) for all-cause mortality comparing MASLD to non-MASLD.

| Number of<br>LTCs | Non-SLD | MASLD | MASLD v non-MALD<br>HR (95%CI) |
| --- | --- | --- | --- |
|  | Events / total | Events / total |  |
| **Overall** |  |  |  |
| 0 | 8955 / 178357 | 5061 / 64823 | 1.27 (1.23, 1.32) |
| 1 | 6361 / 85262 | 4298 / 38312 | 1.19 (1.14, 1.24) |
| 2 | 3488 / 30658 | 2678 / 17112 | 1.09 (1.03, 1.15) |
| 3 | 1584 / 9159 | 1469 / 6581 | 1.09 (1.01, 1.17) |
| 4 | 743 / 2903 | 739 / 2536 | 0.90 (0.81, 1.01) |
| >= 5 | 623 / 1481 | 665 / 1656 | 0.79 (0.71, 0.89) |
| Total | 21754 /<br>307820 | 14910 / 131020 | 1.16 (1.13, 1.19) |
| **Females** |  |  |  |
| 0 | 4530 / 112190 | 1620 / 24056 | 1.38 (1.30, 1.46) |
| 1 | 3386 / 56848 | 1564 / 16979 | 1.29 (1.21, 1.37) |
| 2 | 1821 / 20899 | 995 / 8301 | 1.18 (1.09, 1.28) |
| 3 | 820 / 6200 | 561 / 3362 | 1.12 (1.00, 1.26) |
| 4 | 369 / 1845 | 253 / 1209 | 0.91 (0.77, 1.08) |
| >= 5 | 283 / 883 | 268 / 838 | 0.88 (0.74, 1.05) |
| Total | 11209 /<br>198865 | 5261 / 54745 | 1.25 (1.20, 1.29) |
| **Males** |  |  |  |
| 0 | 4425 / 66167 | 3441 / 40767 | 1.21 (1.16, 1.27) |
| 1 | 2975 / 28414 | 2734 / 21333 | 1.12 (1.06, 1.18) |
| 2 | 1667 / 9759 | 1683 / 8811 | 1.02 (0.95, 1.09) |
| 3 | 764 / 2959 | 908 / 3219 | 1.02 (0.92, 1.13) |
| 4 | 374 / 1058 | 486 / 1327 | 0.91 (0.79, 1.05) |
| >= 5 | 340 / 598 | 397 / 818 | 0.72 (0.61, 0.84) |
| Total | 10545 /<br>108955 | 9649 / 76275 | 1.10 (1.07, 1.13) |

The Cox model was stratified by region and age group, adjusted for sex, ethnicity, education, Townsend Deprivation Index (in fifths), physical activity, smoking status, weekly alcohol consumption, and long-term condition count. For the sex-mixed model, the model was additionally adjusted for sex.

As expected, having more LTCs was associated with increased mortality: compared to people with no LTCs, the HR in those having ≥ 2 LTCs was 2.18 (2.10, 2.27) in the MASLD cohort and 2.42 (2.34, 2.50) in the non-MASLD cohort. In those having ≥ 5 LTCs the HR was 4.26 (3.92, 4.63) and 6.30 (5.80, 6.84), respectively (supplementary figure 2, supplementary table 3). The HR per additional LTC was slightly higher in those without MASLD than in those with (1.38 (1.37, 1.40) vs.1.30 (1.29, 1.31)). Sensitivity analyses excluding the first two years of follow-up and additionally adjusting for the five MASLD-defining cardiometabolic risk factors showed similar results to the primary analysis (supplementary table 4).

Subgroup analyses stratified by age groups, deprivation, education level, smoking status, physical activity and BMI levels also showed consistently positive association between the number of LTCs and all-cause mortality (supplementary figure 3).

### LTCs, cardiometabolic risk factors, and all-cause mortality

In people with MASLD, hypertension, diabetes, and low HDL-cholesterol among all cardiometabolic risk factors were positively associated with all-cause mortality, with HRs 1.09 (1.04, 1.13), 1.46 (1.41, 1.51) and 1.18 (1.14, 1.22), respectively. Only 16 of the 47 prevalent LTCs were associated with increased mortality. These LTCs included heart failure (2.57 (2.06, 3.22)), stroke (1.61 (1.40, 1.85)), heart valve disorder (1.49 (1.22, 1.82)), IHD (1.33 (1.23, 1.43)), arrythmia (1.21 (1.07, 1.38)), chronic kidney disease (CKD) (2.37 (1.91, 2.94)), extrahepatic solid organ cancer (1.70 (1.54, 1.88)), haematological cancers (2.99 (2.29, 3.90)), and chronic obstructive pulmonary disease (COPD) (1.41 (1.23, 1.63)), with Parkinson’s disease showing the highest HR (6.09 (4.47, 8.29)). (table 4)

**Table 4:**
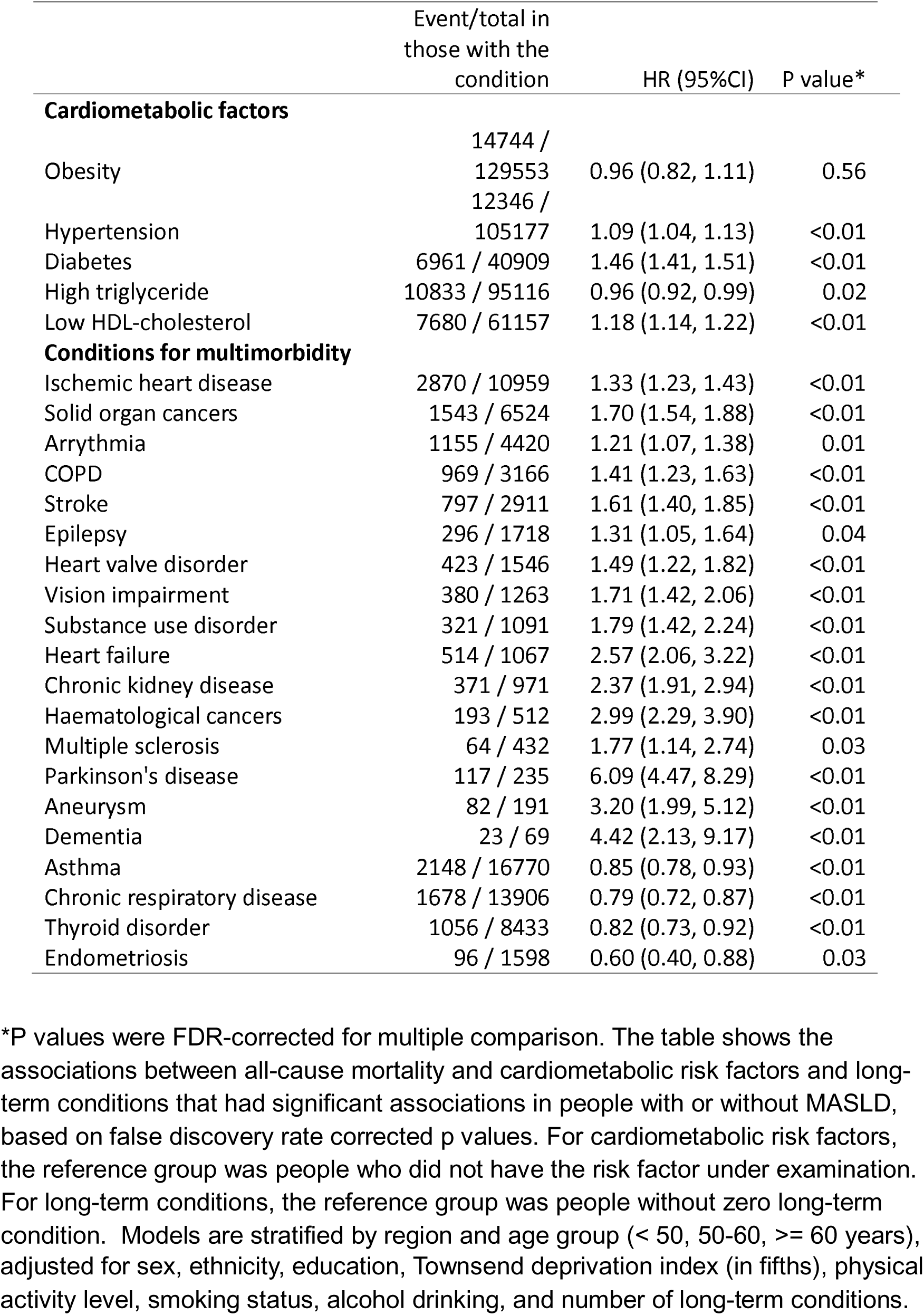
Association between having specific cardiometabolic risk factors and long-term conditions with all-cause mortality in people with MASLD.

## Discussion

This study provides insights into the prevalence and impact of extrahepatic LTCs and multimorbidity among individuals with MASLD compared to those without. Our findings revealed a higher burden of multimorbidity in the MASLD population, particularly with higher prevalences of metabolic, cardiovascular, cancer, renal and mental/behavioural conditions. We also observed positive associations between multimorbidity and all-cause mortality in both cohorts. The association between MASLD and all-cause mortality was dependent on the number of LTCs, with stronger associations in those with lower number of LTCs. LTCs varied in their associations with all-cause mortality. Diabetes, extrahepatic cancers, cardiovascular diseases and CKD showed strong associations, suggesting they should be the target conditions for primary prevention of multimorbidity and optimisation.

MASLD patients exhibited a notably higher prevalence of extrahepatic multimorbidity compared to their non-MASLD counterparts. Specifically, 14.4% of non-MASLD individuals had ≥ 2 LTCs, compared to 21.3% among MASLD individuals, which would further rise to 50.5% if MASLD were counted as an LTC, and to 100.0% if cardiometabolic risk factors were counted. In the non-MASLD cohort, if cardiometabolic risk factors were counted, the prevalence of multimorbidity would rise to 71.6%. We found that MASLD status increased the odds of having a wide spectrum of LTCs, particularly with endocrine/metabolic diseases (including diabetes, gout, thyroid disorder), cardiovascular diseases (hypertension, heart failure, venous thromboembolism, IHD, stroke, etc), extrahepatic cancers, CKD, mental/behavioural disease (PTSD, bipolar disorder, depression, anxiety, etc.) and respiratory diseases (asthma, COPD, chronic respiratory disease, etc.), which aligns with previous research [16,18,34–37].

The mechanisms for these associations may vary from condition to condition, but obesity and the complex interplay between hepatic lipid accumulation and systemic metabolic disturbances provide a foundation for MASLD’s broad associations with multiple LTCs. Insulin resistance, a typical feature of type 2 diabetes, impairs hepatic lipid metabolism, contributing to dysfunction [38]. This metabolic disruption promotes excess lipid accumulation in hepatocytes, chronic liver inflammation, and atherogenic dyslipidaemia. Altered gut microbiota, or gut dysbiosis, and the evolution of certain pathobionts further exacerbate this inflammatory milieu. Moreover, chronic inflammation within the liver extends its impact by activating pathways like NF-κB, reducing adiponectin while increasing leptin levels, which are implicated in the pathogenesis of cancers [39]. Recent genome-wide association analyses have also demonstrated that some underlying genetic variants for MASLD have pleiotropic effects on cardiometabolic and inflammatory traits, including insulin resistance, type 2 diabetes, triglycerides, low density lipoprotein, and obesity [40–42].

We found that MASLD and multimorbidity were both positively associated with all-cause mortality. On average, each additional LTC was associated with 30% and 38% higher mortality in MASLD and non-MASLD populations, respectively, which was similar in males and females, consistent with previous findings [9,11]. Having MASLD was associated with 16% higher mortality, consistent with previous meta-analysis [43] and recent investigations using UK Biobank [4,5]. However, we found that the association between MASLD and mortality was dependent on the LTC counts, with a stronger association in people with lower LTC counts. More specifically, having MASLD was associated with 10-27% higher mortality for people, combining the sexes, with 0 to 3 LTCs, whereas no additional effect on mortality was found for those with 4 LTCs. This may suggest that people with ≥ 4 LTCs experience a saturation effect, where the additive impact of having MASLD diminishes due to already elevated baseline risks, consistent with a previous investigation in the US [44].

The association between MASLD and mortality was stronger in females than in males (HRs 1.25 vs. 1.10), consistently observed within each LTC count. This sex disparity may be attributed to greater differences in prevalences of common and mortality-associated LTCs between MASLD and non-MASLD females, than for males. For example, the ORs were higher in females than in males for IHD, stroke, diabetes, COPD and heart failure, while significantly higher ORs were present with some LTCs in females, but not in males, such as extrahepatic cancers and multiple sclerosis (table 2).

Our analysis highlights that extrahepatic cancers, diabetes, CKD, COPD and cardiovascular diseases (heart failure, IHD, stroke, etc.) significantly contribute to increased mortality risk. This emphasizes the substantial potential for reducing mortality via primary prevention and/or aggressive management of these key co-existing conditions, with large benefit of mortality reduction. The high burden of multimorbidity in people with and without MASLD calls for integrated care approaches that address multiple comorbid conditions simultaneously [45]. Given the progressive nature of MASLD and its close association with metabolic and cardiovascular conditions, routine screening for these comorbidities should be an integral part of MASLD management [46,47]. Lifestyle modifications and risk factor management are recommended public health interventions to reduce the incidence and progression of MASLD, thereby potentially mitigating the broader burden of multimorbidity and associated mortality [47]. The recent introduction of GLP1RAs [48] will likely revolutionize the treatment of MASLD and many of its co-morbid conditions. It will be important to ensure equal access to these powerful medications.

This study provides a more detailed exploration of how multimorbidity differed in people with and without MASLD, and the interactions between MASLD and multimorbidity by considering a wide array of LTCs and their specific contributions to mortality. Nevertheless, several limitations should be acknowledged. First, we used FLI to evaluate steatosis, instead of more accurate imaging techniques, which are the current gold standard. However, previous evidence has shown that FLI yielded high accuracy [5] and was more accurate than other steatosis indices [49], when compared to imaging-based assessment. Second, the UK Biobank population is a predominantly White cohort with higher socioeconomic status and health status, compared to the general UK population; therefore, the prevalence estimates may not be representative of the whole UK population [50]. This may limit the generalizability of our findings to populations of different ethnic or socioeconomic backgrounds. Third, we replied on self-reported data for lifestyle factors, including physical activity, smoking and alcohol consumption, which may be susceptible to information biases. Particularly, inaccuracies in self-reported alcohol consumption may result in misclassification of excessive alcohol drinking, and consequently, affecting the accurate identification of MASLD cases. Fourth, the primary exposure was the number of LTCs at baseline, and future studies are warranted to investigate specific LTC clusters and trajectory patterns.

In conclusion, this study revealed that individuals with MASLD had a substantially higher burden of multimorbidity compared to those without MASLD, particularly in relation to diabetes, extrahepatic cancer, COPD, CKD and cardiovascular diseases. It also elucidated the interplay between MASLD and multimorbidity on all-cause mortality. Having one additional LTC was associated with 30% higher mortality risk, whereas for the same LTC counts, the association between additionally having MASLD and all-cause mortality was more pronounced in those with low number of LTCs. Therefore, addressing multimorbidity in MASLD patients through multidisciplinary and proactive management of multimorbidity is crucial to improving patient outcomes and reducing the overall public health impact of MASLD.

## Data Availability

All data produced are available online at UK Biobank

## Abbreviations

ALD: alcohol-related liver disease
BMI: body mass index
BP: blood pressure
CI: confidence interval
CKD: chronic kidney disease
COPD: chronic obstructive pulmonary disease
FDR: false discovery rate
FLI: fatty liver index
HbA1c: glycated haemoglobin
HDL: low-density lipoprotein
HR: hazard ratio
IHD: ischemic heart disease
LTC: long-term conditions
MASLD: metabolic dysfunction-associated steatotic liver disease
MetALD: metabolic dysfunction and alcohol-related liver disease
PTSD: post-traumatic stress disorder
SD: standard deviation
TG: triglycerides
TIA: transient ischemic attack

## Statements

## Acknowledgement

We sincerely thank the UK Biobank participants and staff for their contribution to this valuable data resource. This study was conducted under application number 74018.

## Data sharing statement

UK Biobank data are available to registered researchers at https://www.ukbiobank.ac.uk/. Analytic codes are available upon request.

## Author contribution

QF and MW conceived the research idea. QF conducted data analysis. QF, AB and MW interpreted results. QF drafted the manuscript. All authors critically reviewed and revised the manuscript.

## Conflict of interests

None.

## Funding

This research/study/project was funded/supported by the NIHR Imperial Biomedical Research Centre (BRC) [NIHR203323]. The views expressed are those of the author(s) and not necessarily those of the NIHR or the Department of Health and Social Care.

AB is supported by National Institutes of Health (NIH)/ National Cancer Institute (NCI) 1U01CA288425-01.

The Section of Investigative Medicine and Endocrinology at Imperial College London is funded by grants from the MRC, NIHR and is supported by the NIHR Biomedical Research Centre Funding Scheme and the NIHR/Imperial Clinical Research Facility. CI is funded by an NIHR Senior Clinical and Practitioner Research Award.

The Division of Digestive Diseases at Imperial College London receives financial support from the National Institute of Health Research (NIHR) Imperial Biomedical Research Centre (BRC) based at Imperial College London and Imperial College Healthcare NHS Trust. BHM is the recipient of a Medical Research Council (MRC) Clinician Scientist award (MR/Z504002/1).

MW is supported by an Australian National Health and Medical Research Council Investigator Grant (APP1174120).

## Disclosure of Ethical Statements

Approval of the research protocol: This study was conducted under application number 74018.

Informed Consent: N/A

Registry and the Registration No. of the study/trial: N/A Animal Studies: N/A

Research involving recombinant DNA: N/A

## Notes

### Competing Interest Statement

The authors have declared no competing interest.

### Funding Statement

This study did not receive any funding

### Author Declarations

The study used (or will use) ONLY openly available human data that were originally located at UK Biobank

